# The treatment gap for mental disorders in adults enrolled in HIV treatment programs in South Africa: A cohort study using linked electronic health records

**DOI:** 10.1101/2020.08.10.20171058

**Authors:** Yann Ruffieux, Orestis Efthimiou, Leigh L. Van den Heuvel, John A. Joska, Morna Cornell, Soraya Seedat, Johannes P. Mouton, Hans Prozesky, Crick Lund, Nicola Maxwell, Mpho Tlali, Catherine Orrell, Mary-Ann Davies, Gary Maartens, Andreas D. Haas

## Abstract

**Background:** Mental disorders are common in people living with HIV (PLWH) but often remain untreated. We aimed to explore the gap in access to treatment (treatment gap) for mental disorders in adults followed-up in antiretroviral therapy (ART) programs in South Africa and disparities in access to mental health services.

**Methods:** We conducted a cohort study using ART program data and linked pharmacy and hospitalization data to estimate the 12-month prevalence of treatment for mental disorders (pharmacological or inpatient) and to examine factors associated with the rate of treatment for mental disorders among adults, aged 15-49 years, followed-up from January 1, 2012 to December 31, 2017 at one private care, two pubic primary care, and one public tertiary care ART programs in South Africa. We calculated the treatment gap for mental disorders as the discrepancy between the 12-month prevalence of mental disorders in PLWH (aged 15-49 years) in South Africa (estimated based on data from the Global Burden of Disease study) and the 12-month prevalence of treatment for mental disorders in ART programs. We calculated adjusted rate ratios (aRR) for factors associated with the rate of treatment of mental disorders using Poisson regression.

**Results:** 182,285 ART patients were followed-up over 405,153 person-years. In 2017, the estimated treatment gap for mental disorders was 40.5% (95% CI 19.5%-52.9%) for patients followed-up in private care, 96.5% (95% CI 95.0%-97.5%) for patients followed-up in public primary care, and 65.0% (95% CI 36.5%-85.1%) for patients followed-up in public tertiary-care ART programs. Rates of treatment with antidepressants, anxiolytics and antipsychotics were 17 (aRR 0.06, 95% CI 0.06-0.07), 50 (aRR 0.02 95% CI 0.01-0.03), and 2.6 (aRR 0.39, 95% CI 0.35-0.43) times lower in public primary-care programs than in the private-sector ART program.

**Interpretation:** There is a large treatment gap for mental disorders in PLWH in South Africa and substantial disparities in access to mental health service between patients receiving ART in the public vs. the private sector. In the public sector and especially in public primary care, PLWH with common mental disorders remain largely untreated.

## Introduction

Mental illness is a leading cause of disease burden in sub-Saharan Africa [1,2]. In South Africa, mental disorders affect one in three adults during their lifetime [3]. The Global Burden of Disease (GBD) study estimates a 12-month prevalence of mental disorders among adults 15-49 years old at 15%. Depression and anxiety disorders are the most common mental disorders in South Africa, each affecting 4-5% of the adult population (aged 15-49 years) [4], Severe mental disorders including schizophrenia, psychosis and bipolar disorder are less common and usually occur in less than 1% of the population [4],

South Africa has the largest number of people living with HIV (PLWH) globally. In 2019 there were 7.7 million PLWH in South Africa, over 5 million of whom were receiving antiretroviral therapy (ART) [5]. Mental disorders are highly prevalent among PLWH [6-8] and are associated with suboptimal HIV treatment outcomes including high rates of loss to follow-up, poor adherence and virological response to ART, and increased mortality [9-11].

Early diagnosis and management of mental disorders not only improves the quality of life of people affected by mental health problems, but may also prevent HIV disease progression, development of drug resistance, and HIV transmission [12]. Despite the impact of mental health at all levels of the HIV treatment cascade, there is a large ‘treatment gap’ for mental disorders in South Africa. The treatment gap refers to the difference between the prevalence of a disorder and the proportion of people who are receiving treatment for this disorder [13].

Estimates of the treatment gap for mental disorders in the general population in low-, middle-, and high-income countries range from 50% to over 90% [13-18]. Previous work on the treatment gap in mental health care has focused on the general population. To the best of our knowledge, there are no data on the treatment gap for mental disorders in PLWH in low- and middle- income countries.

The aims of this study are to quantify the treatment gap for mental disorders among PLWH who enrolled in HIV care at public- and private-sectors ART programs in South Africa and to examine demographic and socioeconomic disparities in access to mental health care.

## Methods

### Study design

We calculated the prevalence of mental disorders in PLWH in South Africa using Global Burden of Disease (GBD) 2017 study estimates for the prevalence of mental disorders in adults (aged 15-49 years) in the general South African population [19] and a literature estimate for excess mental disorders in HIV-positive compared with HIV-negative populations [20]. We estimated the 12-month prevalence of treatment for mental disorders and compared rates of treatment for mental disorders in adults follow-up on ART using routine ART program data (covering the period from January 1, 2004, and December 31, 2017) and linked hospitalization and pharmacy claim data (covering the period from January 1, 2012 to December 31, 2017) from three public-sector and one large private-sector ART programs. We followed patients from baseline (i.e. the date of ART initiation or January 1, 2012, whichever occurred later) to their last visit of the ART clinic or their 50th birthday, whichever occurred first. The Western Cape Provincial Health Data Centre (PHDC) linked data from public-sector ART programs (Gugulethu, Khayelitsha, and Tygerberg Hospital) to hospitalization and pharmacy dispensing records from most public-sector health facilities in the Western Cape Province using unique person identifiers [21], The Aid for AIDS (AfA) program linked ART records of patients who are enrolled in private-sector HIV care to hospitalization and pharmacy claims data from the medical insurance fund claim database [22,23]. The four ART programs participate in the Southern African region of the International epidemiology Databases to Evaluated AIDS (IeDEA-SA) [24],

### Patients

In line with the age groups used in the GBD study, we included people aged 15-49 years at baseline who enrolled in the Gugulethu, Khayelitsha, Tygerberg, or AfA ART programs, and had at least one ART follow-up visit after baseline.

### Treatment programs

The South African health system comprises of a public and a private sector. In 2018, over 70% of the South African households primarily accessed public sector care, although it is estimated that >90% of PLHIV use the public sector for their HIV care. Less than 20% of the population are members of a medical aid scheme and have coverage for health care in the private sector [25]. Tygerberg Academic Hospital, Gugulethu Community Health Centre (CHC), and the Khayelitsha ART program are public-sector HIV programs providing ART according to national treatment guidelines [26]. The three programs are situated in Cape Town in South Africa. Gugulethu and Khayelitsha are public primary-care programs located in townships and Tygerberg is a public tertiary-care facility which manages patients with more severe illness. In Cape Town’s public-sector health care system, primary-care facilities are the first point of care for individuals with both common and stable, serious mental disorders. Individuals with serious mental disorders requiring either admission or more specialized services are referred to and managed at secondary- and tertiary-care facilities [27], AfA is a large private-sector HIV management program for insured and employed people in South Africa. HIV treatment is provided by private medical practitioners and specialists following national treatment guidelines [26]. In the private sector, mental health care is provided by independent general practitioners, psychiatrists and psychologists and private inpatient mental health facilities.

Involuntary admissions (e.g. for persons with severe mental illness, including psychotic disorders) are however handled by state services.

### Measures

In line with the GBD 2017 study, we included the following ICD-10 codes as mental disorder: F04-F06.1, F06.3-F07.0, F08-F09.9, F20-F99, G47-G47.29, G47.4-G47.9, R40-R40.4, R45-R49, Z03.2, Z04.6-Z04.72, Z13.4, Z64, Z81, Z81.8, Z86.5, and Z86.59 [28]. Dementia (F01-F03) and substance use disorders (F10-F19) were not included in this analysis because these conditions are regarded as mental disorders but as separate categories in the GBD study [29]. We defined mental health treatment as either inpatient or pharmacological treatment of a mental disorder. Patients who had been admitted for a mental disorder (as defined above) or to a psychiatric health facility were considered to have received inpatient treatment for a mental disorder. Inpatient mental health treatment was further classified as being for a psychotic disorder (ICD-10 codes F20-F29), mood disorder (F30-F39), or anxiety disorder (F40-F48). Patients who had received antipsychotics (Anatomical Therapeutic Chemical [ATC] code N05A), or anxiolytics (N05B), antidepressants (N06A), or psychostimulants (N06B) or a combination of psychiatric drugs (N06C) were considered to have received pharmacological treatment for a mental disorder. We did not consider non-pharmacological outpatient interventions such as psychotherapy in our analysis.

### Statistical analysis

We calculated the treatment gap δ(y, c) in year y and type of care *c* (private, public primary, or public tertiary), using the following equation:

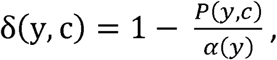

where *P*(*y*, *c*) is the 12-month prevalence of treatment for a mental disorder in year *y* and type of care *c*, and *α(y)* is the prevalence of mental disorders in PLWH in South Africa in year *y*. To estimate the 12-month prevalence of treatment for a mental disorder, *P*(*y*, *c*) we split follow-up time in 12-month intervals, assigned each interval to the calendar year in which most days of the interval occurred and calculated the proportion of patients who had received treatment for a mental disorder (inpatient or pharmacological) during each completed 12-month interval. We did not include follow-up time from incomplete 12-month intervals. For example, if a patient was followed for 4.5 years, then we included the first four 12-month intervals but not the last six months of follow-up. We calculated 95% confidence intervals (CIs) for *P*(*y*, *c*) based on a binomial distribution. We estimated *α*(*y*) based on data from the GBD study [19] and the literature [20] using a set of equations. We provide details on these equations and on the estimation of the 95% CIs for *P*(*y*, *c*) and *δ*(*y*) in the appendix p 23. We expressed δ(y, c) as a percentage throughout our analysis.

We calculated rates of treatment for mental disorders (inpatient, pharmacological or any) for each calendar year as the number of treatment events divided by the number of person-years (py) under follow-up. We treated each hospital admission as a separate event. We treated pharmacy refills for psychiatric medication which occurred in the same year as one event. We did not consider inpatient and pharmacological treatment as mutually exclusive, i.e. a patient who received psychiatric medication while being admitted to a hospital received inpatient and pharmacological mental health treatment simultaneously. We calculated adjusted rate ratios (aRR) for factors associated with the incidence of inpatient treatment of psychotic, mood, anxiety, or any mental disorders, or pharmacological treatment of mental disorders with antipsychotics, antidepressants, anxiolytics, or any psychiatric medication using Poisson regression. We adjusted the Poisson models for type of care (private care [AfA], public primary care [Gugulethu and Khayelitsha], and public tertiary care [Tygerberg]), sex, age (15-24 years, 25-34 years, 35-44 years, 45-49 years), year (2012, 2013, 2014, 2015, 2016, and 2017), and CD4 count at baseline (0-99 cells/μL, 100-199 cells/μL, 200-349 cells/μL, 350-499 cells/μL, 500 cells/μL and over, and missing). Age and year were modelled as time-varying covariates. We used robust sandwich estimators of the standard error to account for clustering of data by patient [30]. We imputed missing data for ICD-10 codes for unclassified hospital admissions (ICD-10 code F99 or missing) using multiple imputation with chained equations [31] and pooled the analyses from 5 sets of imputations using Rubin’s rule [32], We included a dummy variable for missing CD4 cell count information at baseline. Data management and analysis were performed using Stata (Version 15, Stata Corporation, College Station, TX, USA) and the R Project for Statistical Computing Software.

### Ethical considerations

We obtained routine data from the International epidemiology Databases to Evaluate AIDS Southern Africa collaboration (IeDEA-SA) [24], All cohorts participating in IeDEA-SA have ethical approval to examine long-term outcomes of patients through linkage to other datasets, and to contribute de-identified data to the IeDEA-SA Data Centres at the Universities of Cape Town and Bern. The Human Research Ethics Committee of the University of Cape Town, South Africa and the Cantonal Ethics Committee, Bern, Switzerland granted ethical permission for analysis of this database and waived the requirement to obtain informed consent.

## Results

### The prevalence of mental disorders in PLWH in South Africa

We estimated that in 2012, 39.5% (95% CI 29.2-50.0) of PLWH (15-49 years) in South Africa had a mental disorder. The prevalence was 39.4% (29.1-49.8) for 2013, 39.3% (29.0-49.7) for 2014, 39.3% (28.9-49.5) for 2015, 39.2% (29.0-49.4) for 2016, and 39.1% (29.0%-49.3%) for 2017.

### Characteristics of ART patients

A total of 363,384 people were included in the leDEA databases of the Gugulethu, Khayelitsha, Tygerberg and AfA ART programs, of whom 182,285 (50.2%) met our inclusion criteria: 140,322 (77.0%) came from the private-sector AfA program, 39,381 (21.6%) came from the two public primary-care programs (Gugulethu and Khayelitsha) and 2,582 (1.4%) came from the public tertiary-care program (Table 1). Most patients were female (67.5%), median age at ART initiation was 35 years (IQR 31, 41), median CD4 at ART initiation was 238 cells/|aL (IQR 127, 380), and median CD4 at baseline was 359 cells/nL (IQR 219, 548). Patients in the private-sector program initiated ART at an older age and at a higher CD4 cell count than patients in the public-sector programs. CD4 at baseline was higher in private sector than in the public-sector programs.

**TABLE 1.** Characteristics of patients aged 15 year or older under active follow-up in private care, public primary-care, and pub lie tertiary-care antiretroviral therapy programs during 2012-2017^a^

|  | Type of care<sup>b</sup> |  |  |  |
| --- | --- | --- | --- | --- |
|  | Private care<sup>c</sup><br>(n=140322) | Public primary-care<sup>c</sup><br>(n=39381) | Public tertiary-care<sup>c</sup><br>(n=2582) | Total<br>(n=182285) |
| **Characteristics at ART initiation** |  |  |  |  |
| Female, n (%) | 93249 (66.5) | 28076 (71.3) | 1757 (68.0) | 123082 (67.5) |
| Median (IQR) age, years | 36 [32, 41] | 32 [28, 38] | 33 [28, 39] | 35 [31, 41] |
| Median CD4 cells/μL (IQR) | 254 [135, 410] | 193 [107, 302] | 162 [87, 242] | 238 [127, 380] |
| **Characteristics at baseline<sup>d</sup>** |  |  |  |  |
| Median years on ART | 0.3 [0.0, 2.0] | 0.0 [0.0, 2.0] | 1.2 [0.0, 3.7] | 0.1 [0.0, 2.1] |
| Median (IQR) age, years | 38 [33, 43] | 34 [29, 39] | 36 [30, 41] | 37 [32, 42] |
| Median CD4 cells/μL (IQR)<sup>e</sup> | 378 [229, 571] | 307 [184, 454] | 320 [189, 493] | 359 [219, 548] |
| CD4 category<sup>e</sup>, n (%) |  |  |  |  |
| 0-99 cells/μL | 11383 (8.1) | 3381 (8.6) | 265 (10.3) | 15029 (8.2) |
| 100-199 cells/μL | 14075 (10.0) | 4480 (11.4) | 358 (13.9) | 18913 (10.4) |
| 200-349 cells/μL | 30171 (21.5) | 9224 (23.4) | 668 (25.9) | 40063 (22.0) |
| 350-499 cells/μL | 26364 (18.8) | 5864 (14.9) | 456 (17.7) | 32684 (17.9) |
| 500+ cells/μL | 40436 (28.8) | 5519 (14.0) | 563 (21.8) | 46518 (25.5) |
| Missing | 17893 (12.8) | 10913 (27.7) | 272 (10.5) | 29078 (16.0) |

ART: antiretroviral therapy; IQR, interquartile range.
<sup>a</sup> Patients with at least one follow-up visit between January 01, 2012 and December 31, 2017 were included.
<sup>b</sup> All values in the table represent medians and interquartile ranges unless otherwise stated.
<sup>c</sup> Private care, Aid for AIDS program; public primary-care: Guglethu and Khayelitsha programs; public tertiary-care: Tygerberg Hospital.
<sup>d</sup> Baseline was defined as January 01, 2012 or date of ART initiation of the patient, whichever came second.
<sup>e</sup> The CD4 cell count measurement closest to the baseline date, within a 6-month window (before or after)

### The 12-month prevalence of treatment for mental disorders in ART patients

Figure 1 shows the 12-month prevalence of inpatient, pharmacological or any (inpatient or pharmacological) treatment for a mental disorder in private care, public primary care, and public tertiary care ART patients. In private care ART patients, the 12-month prevalence of any treatment for a mental disorder increased from 14.4% (95% CI 14.0%-14.7%) in 2013 to 22.5% (95% CI 22.2%- 22.9%) in 2014 and remained at similar levels between 2015-2017. In public primary care ART patients, the 12-month prevalence of any treatment was 1.0% (95% CI 0.8%-1.2%) in 2012, peaked at 1.9% (95% CI 1.7%-2.1%) in 2015 and declined to 1.4% (95% CI 1.1%-1.7%) in 2017. In public tertiary care ART patients, the 12-month prevalence of any treatment increased gradually from 6.8% (95% CI 4.9%-8.7%) in 2012 to 13.8% (95% CI 5.4%-22.2%) in 2017 (Figure 1). Estimates for the 12-month prevalence of pharmacological treatment for a mental disorder were similar to estimates for the 12-month prevalence of any treatment (Figure 1). In private care, the 12-month prevalence of inpatient treatment for a mental disorder increased from 1.0% (95% CI 0.9%-l.l%) in 2014 to 2.0% (95% CI 1.8%-2.1%) in 2015 and increased further to 2.4% (95% CI 2.2%-2.6%) in 2017 (Figure 1). In public primary care ART patients, the 12-month prevalence of inpatient treatment remained at very low levels throughout the entire study duration, peaking at 0.3% (95% CI 0.2%-0.3%) in 2015. In public tertiary care ART patients the 12-month prevalence of inpatient treatment increased from 0.3% (95% CI 0.0%-0.7%) in 2012 to 1.1% (95% CI 0.0%-2.2%) in 2016 (Figure 1). In the private-sector program the 12-month prevalence of inpatient and pharmacological mental health treatment was higher in women than in men (Figure SI).

**Figure 1.**
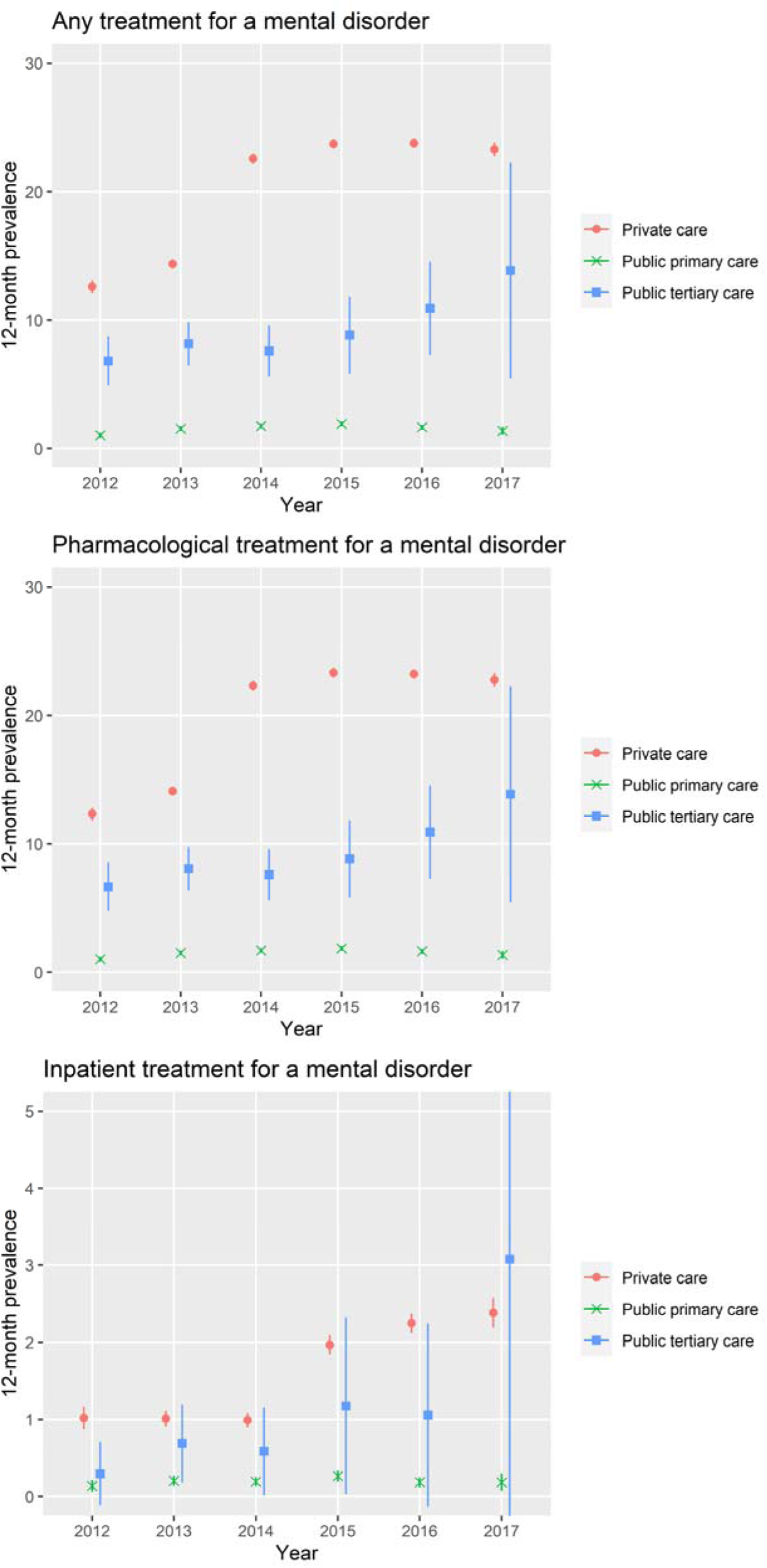
12-month prevalence of treatment for a mental disorder in patients aged 15-49 years followed-up in private care (AfA), public primary-care (Gugulethu, Khayelitsha), and public tertiary-care (Tygerberg) ART programs, 2012-2017. Patients who had been admitted for a mental disorder or to a psychiatric health facility were considered to have received inpatient treatment for a mental disorder. Patients who had received antipsychotics (Anatomical Therapeutic Chemical [ATC] code N05A), anxiolytics (N05B), antidepressants (N06A), psychostimulants (N06B) or psychiatric combination drugs (N06C) were considered to have received pharmacological treatment for a mental disorder. Patient who received either inpatient or pharmacological treatment were considered to have received any treatment for a mantel disorder.

### The mental health treatment gap in PLWH who enrolled in ART programs

Figure 2 and Table S2 show the estimated treatment gap for mental disorders in private care, public primary care, and public tertiary care ART patients. In private care ART patients, the treatment gap decreased from 63.6% (95% CI 50.5%-71.2%) in 2013 to 42.7% (95% CI 22.3%-54.7%) in 2014 and remained at similar levels through the rest of the study period (Figure 2). In public primary care ART patients the treatment gap decreased slightly from 97.4% (95% CI 96.3%-98.1%) in 2012 to 96.5% (95% CI 95.0%-97.5%) in 2017 and in public tertiary care ART patients the treatment gap decreased from 82.8% (95% CI 74.4%-88.4%) in 2012 to 65.0% (95% CI 36.5%-85.1%) in 2017.

**Figure 2.**
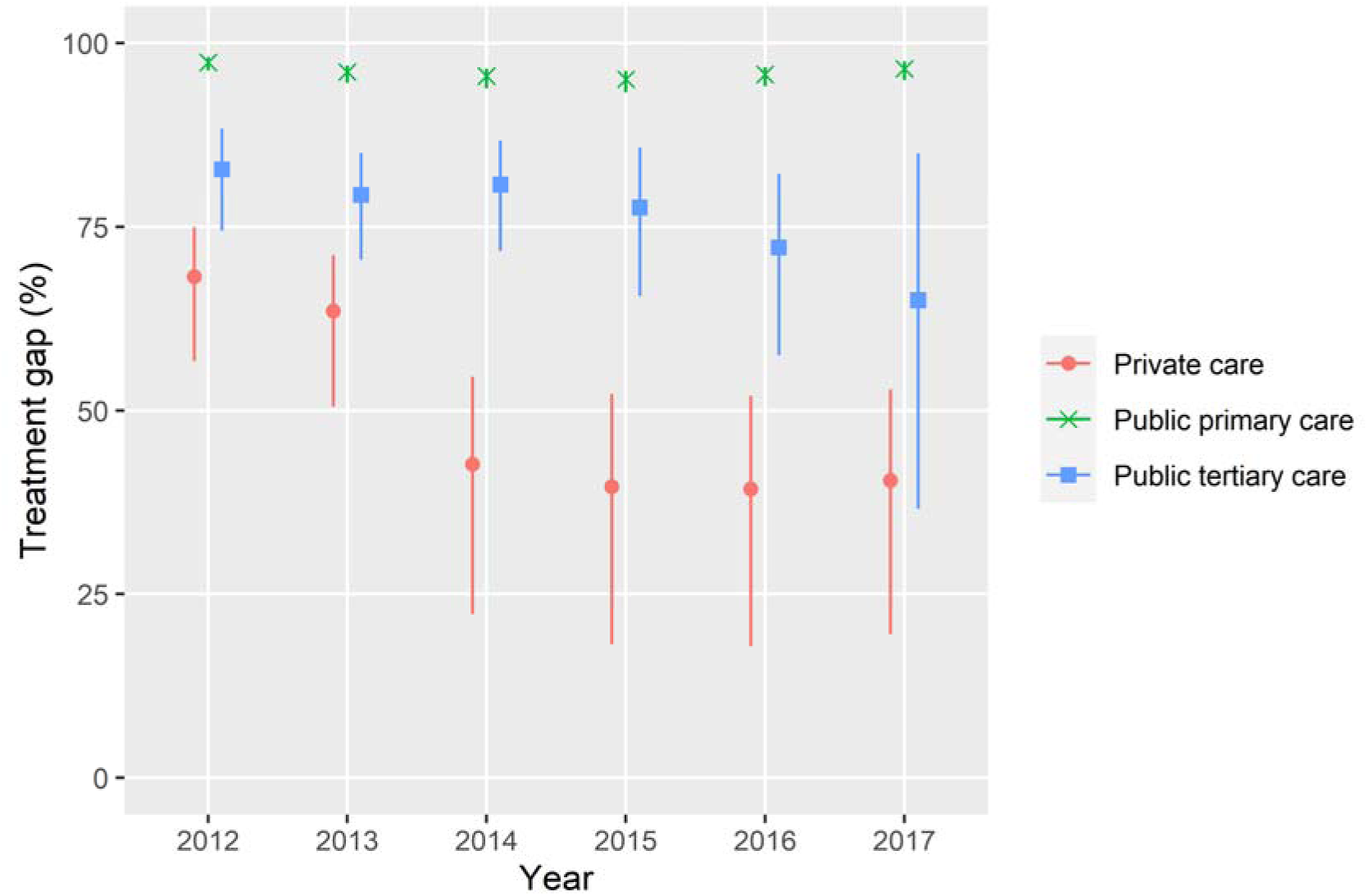
Treatment gap for mental disorders at private care (AfA), public primary-care (Gugulethu, Khayelitsha), and public tertiary-care (Tygerberg) antiretroviral therapy programs, 2012-2017.

### Differences in mental health treatment rates by type of care

In Table SI. we show the number of treatment events, py at risk and crude rates of treatment for mental disorders among patients followed-up in private care, public primary-care, and public tertiary-care ART programs for the years 2012-2017. Figure 3 shows aRRs comparing rates of treatment for mental disorders between patients enrolled in private care, public primary care, and public tertiary care ART programs. The rate of inpatient treatment was 25 times (aRR 0.04, 95% CI 0.03-0.05) lower for mood disorders, 14 times lower (aRR 0.07, 95% CI 0.04-0.14) for anxiety disorders, and about two times higher (aRR 1.8, 95% CI 1.31-2.47) for psychotic disorders in patient enrolled in public primary care compared to private-care ART programs. In patients enrolled in public tertiary care compared to private-care ART programs, the rates of inpatient treatment were five times lower (aRR 0.20, 95% CI 0.11-0.36) and less than two times lower (aRR 0.59, 95% CI 0.22-1.6) for mood and anxiety disorders, but seven times higher (aRR 7.26, 95% CI 3.66-14.41) for psychotic disorders. The rates of treatment with antidepressants, anxiolytics, and antipsychotics were 17 (aRR 0.06, 95% CI 0.06-0.07), 50 (aRR 0.02, 95% CI 0.02-0.03), and 2.6 (aRR 0.38, 95% CI 0.35-0.42) times lower in patients enrolled in public primary care than in private-care ART programs. In patients enrolled in public tertiary-care compared to private care ART programs, the rates of treatment with antidepressants (aRR 0.48, 95% CI 0.4-0.58) and anxiolytics (aRR 0.16, 95% CI 0.12-0.21) were lower and the rate of treatment with antipsychotics was higher (aRR 1.57, 95% CI 1.27-1.93) (Table 2).

**Figure 3:**
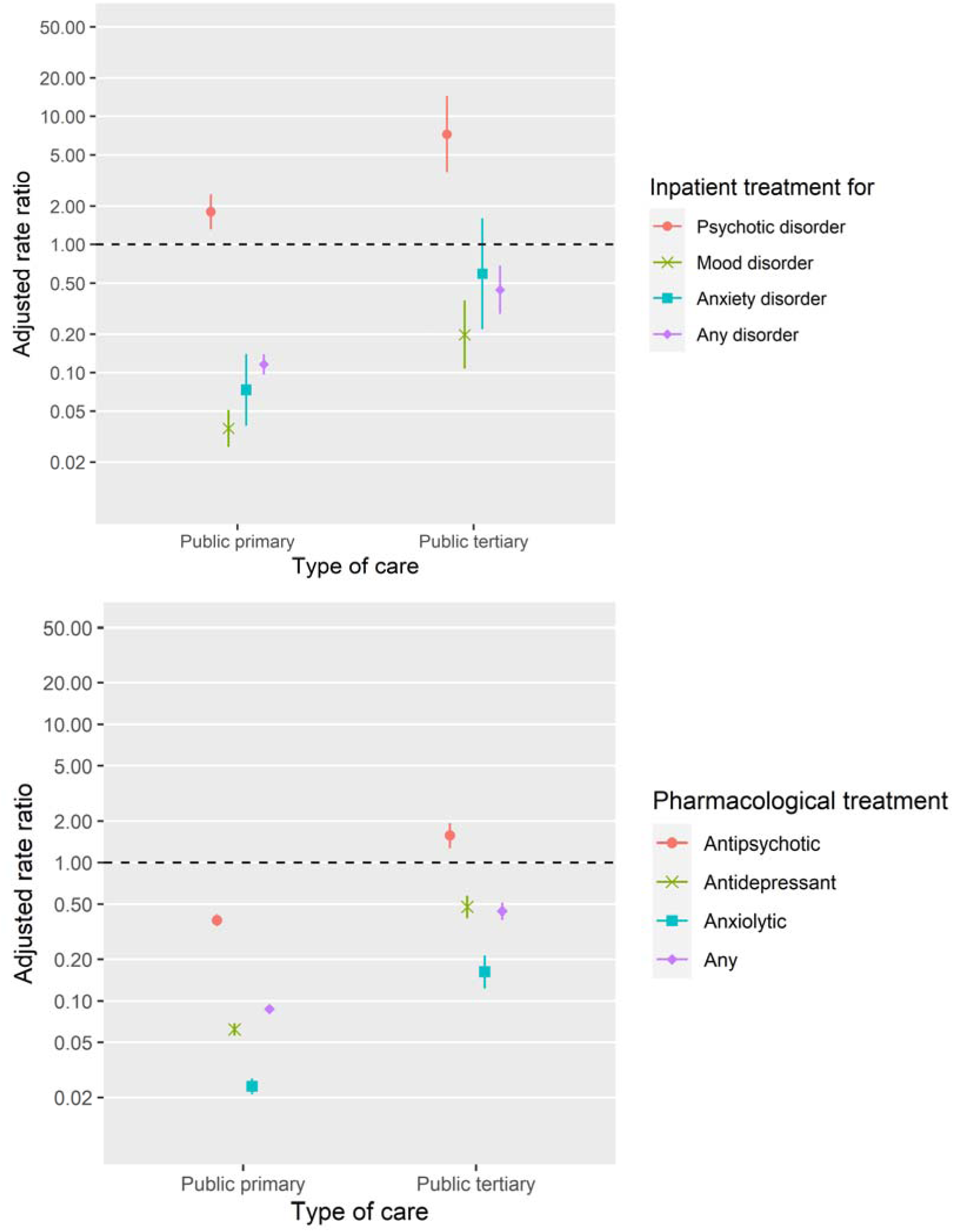
Adjusted incidence rate ratios comparing rates of treatment for mental disorders by type of care Rates of inpatient treatment of psychotic, mood, anxiety, or any mental disorder (top) or pharmacological treatment with antipsychotics, antidepressants, anxiolytics, or any psychiatric medication are compared between patients follow-up at the two public primary-care programs (Gugulethu and Khayelitsha), the public tertiary-care program Tygerberg, and the private care program Aid for AIDS (AfA). The private-care program was the reference group. Incidence rate ratios were adjusted for gender, current age, current year, and baseline CD4 cell count.

**TABLE 2.**
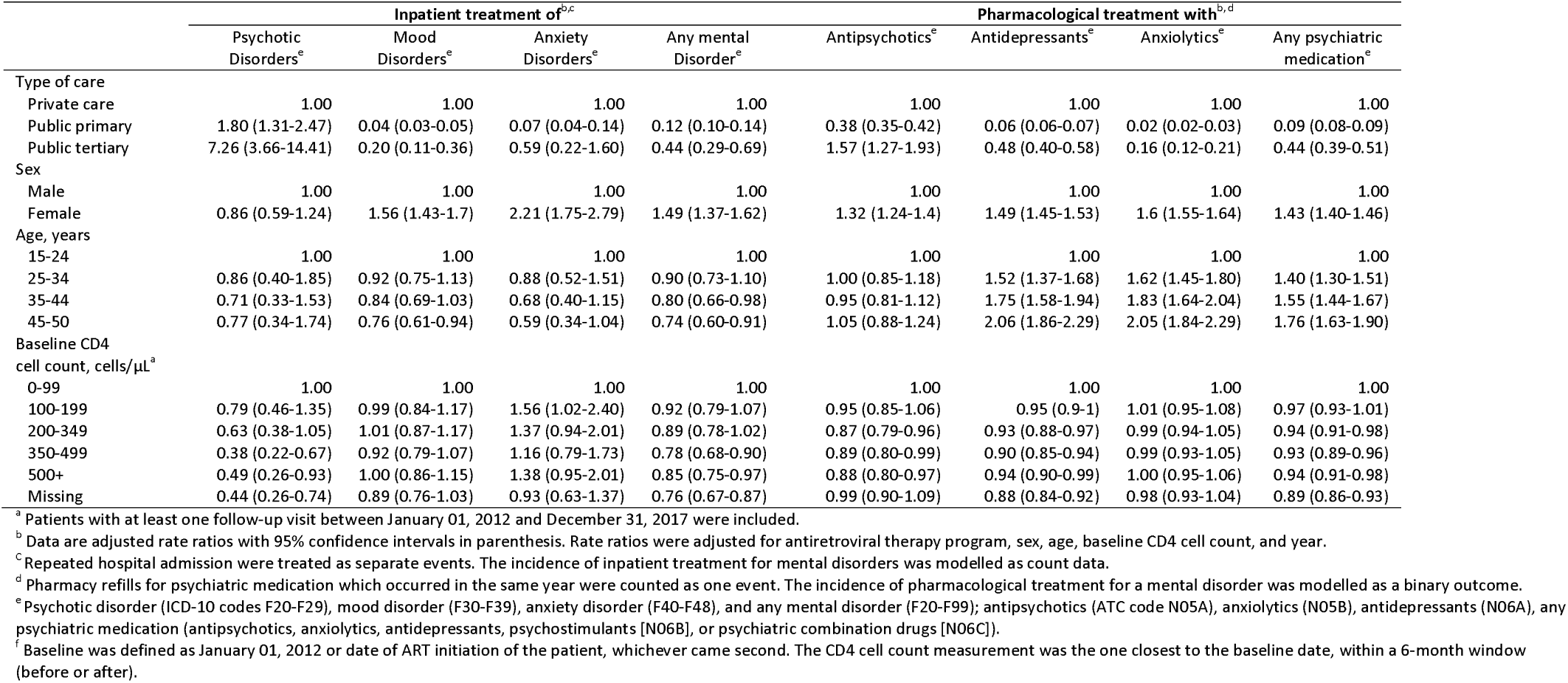
Adjusted incidence rate ratios for factors associated with treatment for mental disorders in patients aged 15-49 year followed-up in private care, public primary-care, and public tertiary-care antiretroviral therapy programs during 2012-2017^a^

### Patient characteristics associated with mental health treatment rates

Women had a higher incidence of inpatient treatment for mood (aRR 1.56, 95%CI 1.43-1.70), anxiety (2.21, 95% CI 1.75-2.79), and any mental disorder (1.49 95% CI 1.37-1.62) than men (Table 2). Women were also more likely to receive psychiatric medication than men with aRRs of 1.32 (95% CI 1.24-1.40) for antipsychotics, 1.49 (95% CI 1.45-1.53) for antidepressants, and 1.60 (95% CI 1.55-1.64) for anxiolytics. Older age was associated with a lower incidence of inpatient treatment for any mental disorders and a higher incidence of pharmacological treatment with antidepressants, anxiolytics, and any psychiatric medication. Higher baseline CD4 cell counts were associated with a lower incidence of inpatient treatment for a psychotic disorder, and with a slightly lower incidence of treatment with antipsychotics and antidepressants (Table 2).

## Discussion

Our study showed a large treatment gap for mental disorders in PLWH who enrolled in ART programs in South Africa. We found substantial disparities in access to mental health services between patients who receive ART in private- vs. in public-sector programs. In 2017 the treatment gap for mental disorders in PLWH was 96.5% in public primary-care programs, 65.0% in the public tertiary-care program, and 40.5% in the private ART program. The rate of treatment with antidepressants, anxiolytics, and antipsychotics was 17, 50, and 2.6 times lower in the public primary-care programs compared with the private program. We found considerable gender differences in mental health treatment rates, with women being treated more often than men.

Our estimates for the treatment gap in mental disorders in PLWH in South Africa are largely consistent with data from earlier studies in the general population from various settings. A meta-analysis of studies published between 1980 and 2003 estimated that on average the global treatment gap was 78% for alcohol abuse and dependence, 58% for generalized anxiety and obsessive compulsive disorders, 56% for depression, dysthymia, and panic disorder, 50.2% for bipolar disorder, and 32% for schizophrenia [13]. The WHO World Mental Health Survey, conducted between 2001 and 2003 in 14 countries, reported a treatment gap for mental health and substance use problems of 76% in less-developed countries and 85% in developed countries [15]. In Nigeria, the only African country included in the survey, the treatment gap was 99.2% [15]. In rural Ethiopia, between 1998 and 2001 the treatment gap was about 90% and 70% for schizophrenia and major depression [16,33]. Similarly, the South African Stress and Health Study (SASH), conducted between 2002 and 2004, found that 74% of the people with anxiety disorder, mood disorder, substance use disorder, or intermittent explosive disorder received no mental health treatment in the year before the survey [17]. Our estimates are also consistent with more recent results from Docrat and colleagues, who estimated the treatment gap for mental disorders in the general South African population based on data on mental health care expenditure and GBD estimates. The study estimated a 92% treatment gap in South Africa’s public sector, with 86% of the mental health care budget spent on inpatient care and only 8% in the primary-care setting [18]. The authors concluded that South Africa’s public health care system focuses on treating the most severe conditions, while early treatment of non-severe conditions or prevention are not priorities [34], The high incidence of hospital admission for psychotic disorders and the low rates of prescriptions of antidepressants and anxiolytics in the primary-care setting provides further support for this conclusion.

The comparison of the rate of inpatient treatments between public- and private-sector programs has to be interpreted with caution. According to the South African Mental Health Care Act all involuntary admissions are handled by state services [35]. Our data did not capture involuntary admissions of private care patients because we could not link records from private care patients to public service data. Because admissions for psychotic disorder are often involuntary [36,37] excess rates of inpatient care for psychotic disorders in patients enrolled in public compared to private-sector ART programs are likely to be overestimated. The large increase of inpatient care for psychotic disorders in patients enrolled in public tertiary care may also reflect more advanced HIV disease in these patients. Psychotic disorders secondary to HIV tend to occur at more advanced stages and the lower baseline CD4 cell counts observed in public, particularly tertiary care, programs could align with this explanation [38]. Furthermore, higher baseline CD4 counts were associated with lower rates of treatment (inpatient and antipsychotics) for psychotic disorders. The higher inpatient treatment rates for anxiety and mood, but not psychotic, disorders in women could represent higher prevalence of common mental disorders in women [39,40]. Other explanations for observed gender differences in treatment rates are differences in health care seeking behavior between men and women or discrimination against men in the health care system, for example through practitioner’s subconscious tendency to overlook psychological distress in men [41]. Lower rates of inpatient treatment and higher rates of pharmacological treatment, excluding antipsychotics, in older age groups could potentially be explained by younger individuals presenting with first episodes of illness requiring more intensive investigations and interventions as compared to individuals with known or recurring conditions.

Our results must be considered in the light of several limitations. Despite a thorough search, we could not find estimates for the overall prevalence of psychiatric disorders in PLWH in South Africa. We therefore estimated the need for mental health treatment based on data from the GBD [19] and literature estimates [20]. Population-based surveys using structured diagnostic interviews [13-17,33] provide more reliable estimates for the need of mental health treatment than our study. We had no data on non-pharmacological outpatient interventions such as psychotherapy and could not consider these therapies when estimating the treatment gap. Considering poor access to psychological interventions (particularly in public services), we believe that this limitation could have led to a slight over-estimation of the treatment gap. Conversely, psychiatric medication is sometimes used for indications not related to mental health, for which reason we might slightly underestimate the treatment gap.

Furthermore, we did not adjust our estimates for the prevalence of mental disorders in PLHIV (i.e. the denominator of the treatment gap) for underlying differences in the prevalence of mental disorders between different care settings. Mental disorders might be more common in public sector patients who tend to have more socio-economic stressors (including unemployment, living in environments where they are exposed to violence and trauma) making them more vulnerable to mental disorders. Patients in tertiary care are likely to have substantially more comorbidities and thus higher risk of mental disorders. Because we could not account for these underling differences in the prevalence of mental disorders, we may have overestimated the treatment gap in private care and underestimated the treatment gap in tertiary care. High prescription rates of psychiatric medication (particularly anxiolytics) in the private care setting could also reflect over-treatment.

An important strength of our study is that we used medical records to estimate mental health treatment utilization rates. Most previous studies on the treatment gap relied on self-report data [13-17,33]. Users of mental health services might underreport service utilization due to mental health stigma [42,43] and such underreporting might lead to overestimation of the treatment gap. A further strength of our study is that we could account for mental health treatment received outside the HIV treatment setting because we linked public- and private sector ART program data to province-wide and private- data on mental health treatment, respectively. Lastly, inclusion of data from public primary-care, public tertiary-care, and private care settings adds to the robustness and generalizability of our findings.

The integration of evidence-based interventions for diagnosing and managing mental disorders in primary-care ART programs holds great promise for closing the treatment gap in people living with HIV [44], This could lead to an improvement of outcomes across the treatment cascade and of long-term retention in care. The World Health Organization and the South African government have published guidelines for the management of mental disorders in non-specialized settings [45,46]. Our study suggests that implementation of mental health services in primary-care ART programs is inconsistent. Continued efforts to close the treatment gap for people with mental disorders are needed. Strategies to strengthen task-shifting, training and capacitation of primary health care staff, and models of referral and stepped-care will be crucial to closing the treatment gap [47,48].

## Conclusion

There is a large treatment gap for mental disorders in people living with HIV in South Africa and substantial disparities in access to mental health service between patients receiving care in the public vs. the private sector. In the public sector and especially in public primary care, PLWH with common mental disorders remain largely untreated.

### Competing interests

The authors declare no competing interests.

### Authors’ contributions

AH and YR wrote the first draft of the study protocol which was revised by OE, LvH, JJ, JM, CL, and GM. All authors reviewed and approved the final version of the study protocol. MC, JM, HP, NM, MT, CO, MD, GM, and AH contributed to data collection. YR and OE did statistical analysis. YR, OE, LvH, JJ, MC, SS, JM, HP, CO, MD, GM and AH interpreted the results. YR and AH wrote the first draft of the manuscript which was revised by all authors. All authors have reviewed and approved the final version of the manuscript.

## Data Availability

All data were obtained through the International epidemiology Database to Evaluate AIDS Southern Africa collaboration (IeDEA-SA, https://www.iedea-sa.org/). Data cannot be made available online because of legal and ethical restrictions. To request data, readers may contact IeDEA for consideration by filling out the online form available at https://www.iedea-sa.org/contact-us/

## Acknowledgements

The study was supported by the US National Institutes of Health (the National Institute of Allergy and Infectious Diseases, the Eunice Kennedy Shriver National Institute of Child Health and Human Development, the National Cancer Institute, the National Institute of Mental Health, the National Institute on Drug Abuse, the National Heart, Lung, and Blood Institute, the National Institute on Alcohol Abuse and Alcoholism, the National Institute of Diabetes and Digestive and Kidney Diseases, and the Fogarty International Center (grant number U01AI069924), the Swiss National Science Foundation (SNSF) (grant numbers 174281, P2BEP3_178602,180083) and the the South African Medical Research Council (SAMRC). The findings and conclusions in this report are those of the authors and do not necessarily represent the official position of the funding agencies.

We thank the Western Cape Provincial Health Data Centre for compiling and providing public sector mental health data and Nicki Tiffin, PhD, MPH for assisting with data compilation.

## Supplemental material

**Figure S1.**
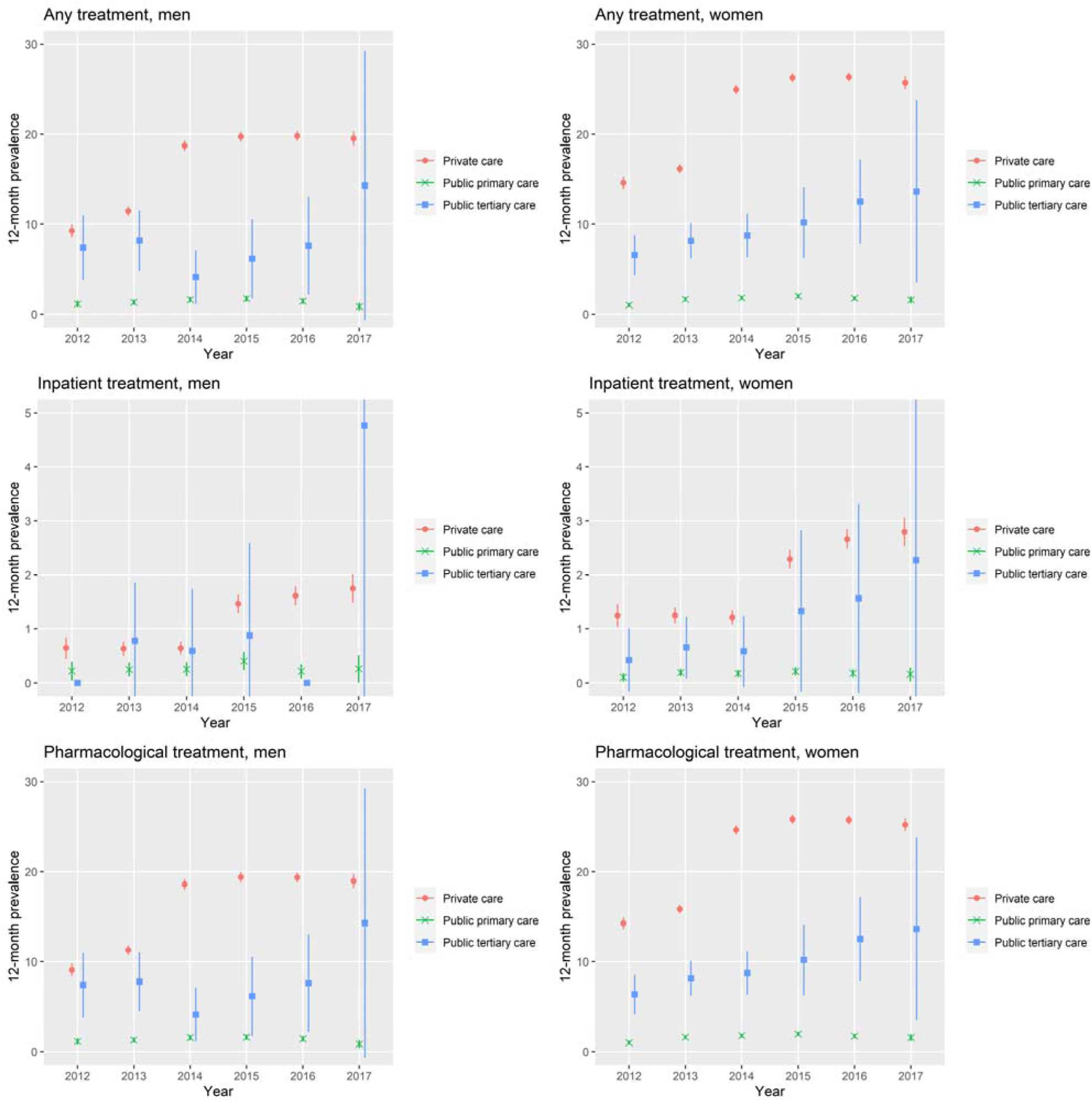
12-month prevalence of treatment for a mental disorder in men and women aged 15-49 years who were followed-up in private care (AfA), public primary-care (Gugulethu, Khayelitsha), and public tertiary-care (Tygerberg) ART programs, 2012-2017. Patients who had been admitted for a mental disorder or to a psychiatric health facility were considered to have received inpatient treatment for a mental disorder. Patients who had received antipsychotics (Anatomical Therapeutic Chemical [ATC] code N05A), anxiolytics (N05B), antidepressants (N06A), psychostimulants (N06B) or psychiatric combination drugs (N06C) were considered to have received pharmacological treatment for a mental disorder. Patient who received either inpatient or pharmacological treatment were considered to have received any treatment for a mantel disorder.

**Table S1:** Rates of treatment for mental disorders (per 100 person years) among patients followed-up in private care (AfA), public primary-care (Gugulethu, Khayelitsha), and public tertiary-care (Tygerberg) ART programs, 2012-2017

| Treatment | Calendar year | Type of care |  |  |  |  |  |  |  |  |
| --- | --- | --- | --- | --- | --- | --- | --- | --- | --- | --- |
|  |  | Private care<sup>a</sup> |  |  | Public primary care<sup>a</sup> |  |  | Public tertiary care<sup>a</sup> |  |  |
|  |  | n<sup>b</sup> | N<sup>c</sup> | Rate (%)<sup>d</sup><br>(95% CI) | n<sup>b</sup> | N<sup>c</sup> | Rate (%)<sup>d</sup><br>(95% CI) | n<sup>b</sup> | N<sup>c</sup> | Rate (%)<sup>d</sup><br>(95% CI) |
| Inpatient | 2012 | 1129 (1410) | 61172 | 1.8 (1.7-2.0) | 33 (43) | 17846 | 0.2 (0.1-0.2) | 7 (11) | 1625 | 0.4 (0.1-0.7) |
|  | 2013 | 393 (512) | 37582 | 1.0 (0.9-1.1) | 46 (55) | 19675 | 0.2 (0.2-0.3) | 8 (13) | 1231 | 0.6 (0.2-1.1) |
|  | 2014 | 411 (532) | 42679 | 1.0 (0.9-1.1) | 49 (60) | 21068 | 0.2 (0.2-0.3) | 8 (11) | 945 | 0.8 (0.3-1.4) |
|  | 2015 | 1076 (1353) | 47223 | 2.3 (2.1-2.4) | 66 (91) | 21316 | 0.3 (0.2-0.4) | 2 (2) | 505 | 0.4 (-0.2-0.9) |
|  | 2016 | 1216 (1630) | 49917 | 2.4 (2.3-2.6) | 35 (42) | 18284 | 0.2 (0.1-0.3) | 4 (4) | 316 | 1.3 (0-2.5) |
|  | 2017 | 1279 (1695) | 49802 | 2.6 (2.4-2.7) | 25 (36) | 13788 | 0.2 (0.1-0.3) | 3 (3) | 180 | 1.7 (-0.2-3.5) |
| Pharmacological | 2012 | 13859 | 61172 | 22.7 (22.3-23) | 191 | 17846 | 1.1 (0.9-1.2) | 134 | 1625 | 8.2 (6.9-9.8) |
|  | 2013 | 4732 | 37582 | 12.6 (12.2-13) | 344 | 19675 | 1.7 (1.6-1.9) | 100 | 1231 | 8.1 (6.6-9.9) |
|  | 2014 | 9894 | 42679 | 23.2 (22.7-23.6) | 420 | 21068 | 2.0 (1.8-2.2) | 93 | 945 | 9.8 (7.9-12.1) |
|  | 2015 | 11410 | 47223 | 24.2 (23.7-24.6) | 461 | 21316 | 2.0 (2-2.4) | 57 | 505 | 11.3 (8.5-14.6) |
|  | 2016 | 11385 | 49917 | 22.8 (22.4-23.2) | 368 | 18284 | 2.0 (1.8-2.2) | 37 | 316 | 11.7 (8.2-16.1) |
|  | 2017 | 11542 | 49802 | 23.2 (22.8-23.6) | 298 | 13788 | 2.2 (1.9-2.4) | 21 | 180 | 11.6 (7.2-17.8) |
| Inpatient or pharmacological | 2012 | 14147 | 61172 | 23.1 (22.7-23.5) | 195 | 17846 | 1.1 (0.9-1.3) | 134 | 1625 | 8.2 (6.9-9.8) |
|  | 2013 | 4816 | 37582 | 12.8 (12.5-13.2) | 354 | 19675 | 1.0 (1.6-2) | 100 | 1231 | 8.1 (6.6-9.9) |
|  | 2014 | 10010 | 42679 | 23.5 (23-23.9) | 431 | 21068 | 2.0 (1.9-2.2) | 93 | 945 | 9.8 (7.9-12.1) |
|  | 2015 | 11616 | 47223 | 24.6 (24.2-25) | 474 | 21316 | 2.2 (2.0-2.4) | 57 | 505 | 11.3 (8.5-14.6) |
|  | 2016 | 11714 | 49917 | 23.5 (23-23.9) | 377 | 18284 | 2.1 (1.9-2.3) | 37 | 316 | 11.7 (8.2-16.1) |
|  | 2017 | 11910 | 49802 | 23.9 (23.5-24.3) | 299 | 13788 | 2.2 (1.9-2.4) | 21 | 180 | 11.6 (7.2-17.8) |

<sup>a</sup> Private care, Aid for AIDS program; public primary care: Gugulethu and Khayelitsha programs; public tertiary-care: Tygerberg Hospital.
<sup>b</sup> Number of patients who received treatment in year. For inpatient treatments the total number of treatment events (accounting for repeated admissions of a patient in the same year) is shown in parenthesis.
<sup>c</sup> Number of person-years (PY) under follow-up in antiretroviral therapy programs in year.
<sup>d</sup> Rate per 100 person-years with 95% confidence intervals (CIs) in parentheses.

**Table S2:** The treatment gap for mental disorders at private care (AfA), public primary-care (Gugulethu, Khayelitsha), and public tertiary-care (Tygerberg) antiretroviral therapy programs, 2012-2017.

| Calendar year | Type of care |  |  |
| --- | --- | --- | --- |
|  | Private care | Public primary care | Public tertiary care |
| 2012 | 68.2 (56.7-74.9) | 97.4 (96.3-98.1) | 82.8 (74.4-88.4) |
| 2013 | 63.6 (50.5-71.2) | 96.1 (94.6-97.0) | 79.4 (70.5-85.1) |
| 2014 | 42.7 (22.3-54.7) | 95.5 (93.9-96.5) | 80.7 (71.6-86.7) |
| 2015 | 39.6 (18.4-52.2) | 95.1 (93.3-96.2) | 77.6 (65.5-85.8) |
| 2016 | 39.3 (18.0-51.9) | 95.7 (94.1-96.7) | 72.2 (57.5-82.2) |
| 2017 | 40.5 (19.5-52.9) | 96.5 (95.0-97.5) | 65.0 (36.5-85.1) |

Data are percentages, with 95% confidence intervals in parentheses.

APPENDIX 1: Estimation of the prevalence of mental disorders (MD) in people living with HIV (PLWH) and the treatment gap for MD, in a given calendar year.

### Estimation of the prevalence of MD in PLWH

The target quantity of interest is the prevalence of MD in PLWH *α* = *P*(*MD* +| *HIV*+), calculated as the probability of having a mental disorder conditional on being HIV-positive in a given year. Similarly, we define *β* = *P*(*MD* +| *HIV−)* as the prevalence of MD in HIV-negative people. We have data on the following quantities, with associated 95% confidence intervals (CI):

- the prevalence of MD in the general population in the given year, *p_m_ = P(MD+)*
- the prevalence of MD in the general population in the given year, *p_h_ = P(MD+)*
- the odds ratio for the prevalence of mental disorders in HIV-positive compared to HIV-positive populations

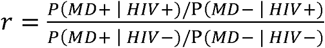

We assume that r ≠ 1.

We obtain the following system of two equations, which we will solve for *a* and *β;*

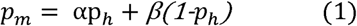

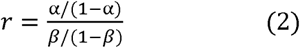

From (1) we find that *β* = (*p_m_* − *αp_h_*)/ (1 − *p_h_*). Inserting this into (2), we obtain:

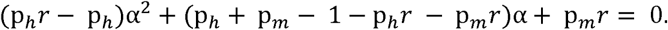

Our desired *α* is a solution to this second degree equation. Setting *α =* (p*_h_r −* p*_h_), b =* (p*_h_ +* p*_m_* − 1 − p*_h_r −* p*_m_r*), *c =* p*_m_r*, and Δ = b^2^ − 4*ac*, the solution in question is

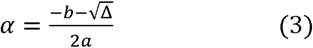

### Estimation of the treatment gap for MD and 95% CI

For a given year, the treatment gap for MD in PLWH followed-up on antiretroviral therapy is estimated as

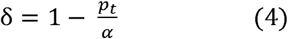

where p*_t_* is the 12-month prevalence of treatment for MD that year. To create a 95% CI for *α*, we assume that p*_m_* and p*_h_* follow Beta distributions, with parameters calibrated so that the 2.5%- and 97.5%-quantiles of the distributions match the bounds of their respective 95% CIs. We assume that log(*r*) follows a normal distribution, with similarly calibrated mean and variance, and that p*_t_* follows a binominal distribution Binom(*p*, *N)* where *p* is the observed 12-month prevalence of treatment for MD and *N* the number of patients at risk.

We independently simulated a large sample of values for p*_m_*, p*_h_, r*, and p*_t_* based on the distributions described above, thereby generating a sample of values for *α* using (3) and δ using (4). The bounds of the 95% CI for 5 are estimated as the 2.5%- and 97.5%-quantiles of the sample for δ.

#### R code to compute confidence intervals for the prevalence of MD in PLWH and for the treatment gap

*#Clb_hiv, CIu_hiv:* lower and upper bounds of 95% confidence interval [Cl) for HIV prevalence in given

#year [provided by GBD)

*#CIb_md, CIu_md:* lower and upper bounds of 95% CI for prevalence of mental disorders in thatyear

#[provided by GBD)

#finding Beta distributions with lower and upper 2.5%-quantiles matching as closely as possible the

#bounds of the 95% CIs for the prevalence estimates provided by GBD

*beta_fitter <- function(par,Clb,Clu) abs(pbeta(CIb,par[1],par[2])-0.025)+abs(pbeta(CIu,par[1],par[2])-0.975) res_hiv <- optim(par=c(l,l),fn=beta_fitter,Clb=Clb_hiv,Clu=Clu_hiv,method=“L-BFGS-B”, lower=c(0.01,0.01), upper=c(I rtf,¡rtf))*

*res_md <- optim(par=c(l,l),fn=beta_fitter,Clb=Clb_md,Clu=Clu_md,method=“L-BFGS-B”, lower=c(0.01,0.01), upper=c(I rtf,¡rtf))*

#parameters of Beta distribution for HIV prevalence *a_hiv <- res_hiv$par[l]*

*b_hiv <- res_hiv$par[2]*

#parameters of Beta distribution for prevalence of mental disorders *ajnd <- res_md$par[l]*

*b_md <- res_md$par[2]*

#generating sample of 100,000 treatment gap estimates

*#logOR, logORyar*. log-odds ratio [and variance thereof) for mental disorders in

#people with HIV vs. people without HIV

*#P:* prevalence of treatment for mental disorders in given year [obtained from ART programs data)

*#N:* total number of persons at risk in given year [obtained from ART programs data)

#function computing point estimate of MD prevalence in PLWH *cond_prev <- function(p_m, p_h, r)*

*{*

*A <- p_h*r-p_h*

*B <- p_h+p_m-l-r*p_h-r*p_m*

*C <- r*p_m*

*delta <- B^A^2-4*A*C*

*(-B-sqrt(delta))/(2*A)*

*}*

*p_h <- rbeta(100000, a_hiv, b_hiv)* # sample for HIV prevalence

*pjn <- rbeta(100000, a_md, b_md)* # sample for prevalence of mental disorders

*r <- exp(rnorm(n=100000, mean=logOR, sd=sqrt(logOR_var))* # sample of odds ratios

*alpha <- cond_prev(p_m, p_h, r)* # sample of estimates for prevalence of mental disorders in PLWH

*p_t<- rbinom(100000, size=N, prob=P)/N* # sample of estimates for prevalence of treatment for MD

*delta <-1 - p_t/alpha* # sample of estimates for the treatment gap

\# 95% CI for prevalence of mental disorders in people with HIV *prevjcl <- quantile(alpha, probs=0.025)*

*prev_ucl <- quantile(alpha, probs=0.025)*

\# 95% CI for treatment gap

*tgapjcl <- quantile(delta, probs=0.025) tgap_ucl <- quantile(delta, probs=0.975)*

## Notes

### Competing Interest Statement

The authors have declared no competing interest.

### Author Declarations

We obtained routine data from the International epidemiology Databases to Evaluate AIDS Southern Africa collaboration (IeDEA-SA) [23]. All cohorts participating in IeDEA-SA have ethical approval to examine long-term outcomes of patients through linkage to other datasets, and to contribute de-identified data to the IeDEA-SA Data Centres at the Universities of Cape Town and Bern. The Human Research Ethics Committee of the University of Cape Town, South Africa and the Cantonal Ethics Committee, Bern, Switzerland granted ethical permission for analysis of this database and waived the requirement to obtain informed consent.

